# Determinants of Repetitive Transcranial Magnetic Stimulation Efficacy in Tobacco Use Disorder: A Pre-Registered Study

**DOI:** 10.64898/2026.06.23.26356059

**Authors:** Michael R. Apostol, Timothy Jordan, Gino Haase, Lucina Q. Uddin, Andrew F. Leuchter, Nicole Petersen

## Abstract

Repetitive Transcranial Magnetic Stimulation (rTMS) is a promising treatment for tobacco use disorder (TUD). Although at a group level, active stimulation outperforms sham, at an individual level, variability exists in clinical response. The behavioral and neurobiological factors that differentiate those who respond to rTMS from those who do not remain unclear. To explore individual factors that influence acute responses to rTMS, *N* = 60 human participants received one session of rTMS to the dorsolateral prefrontal cortex (DLPFC) and to a control region (visual cortex; V5) in a randomized order. They completed behavioral assessments and neuroimaging before and after rTMS sessions. Hypotheses involving behavioral and neuroimaging predictors of response were pre-registered prior to completion of data collection. rTMS to the DLPFC led to significant reductions in self-reported cigarette craving compared with rTMS to a control brain region (*p* = 0.0006) and participants were classified as *n* = 38 responders and *n* = 22 nonresponders. Responders used significantly more cigarettes per day (*M* = 11.441) compared to nonresponders (*M* = 7.952), reported higher levels of cigarette craving (*d* = 1.059), and more severe nicotine withdrawal (*d* = 0.803) prior to rTMS. Neuroimaging analyses based on preregistered hypotheses indicated that DLPFC-frontoparietal and insula whole-brain functional connectivity did not differ significantly between responders and nonresponders. However, exploratory analyses revealed that responders had reduced pre-rTMS functional connectivity between the insula and nucleus accumbens, precuneus, and occipital pole. These findings suggest that response to rTMS for TUD is associated with greater baseline cigarette consumption, craving, and withdrawal, in addition to distinct functional connectivity patterns related to salience, reward, and self-referential processes, providing candidate behavioral and neural markers for personalized rTMS interventions for TUD.

## Introduction

Cigarette smoking is the leading cause of preventable disease and death in the world (Benowitz & Liakoni, 2022; Vellappally et al., 2007; WHO, 2019). Two in three people who smoke want to quit, and most have tried to quit, yet smoking cessation rates are approximately 7% without medical or psychosocial interventions (Babb et al., 2017). Existing interventions include electronic cigarettes, varenicline, bupropion, nicotine replacement therapies, psychotherapies, and community-based interventions (Singh et al., 2026). Among these interventions, varenicline, a partial agonist of α4β2 nicotinic acetylcholine receptors, has shown the greatest efficacy with reported abstinence rates of 18 - 30% (Jiménez-Ruiz et al., 2009; C. J. Jordan & Xi, 2018). Despite this, varenicline and other existing smoking cessation interventions display variable and limited success rates, including the fact that many smokers do not engage with existing interventions, suggesting the need for novel therapeutics to help people quit smoking (Bryant et al., 2011; Caponnetto et al., 2012; Ebbert et al., 2010; Gómez-Coronado et al., 2018; Hartmann-Boyce et al., 2018; Koegelenberg et al., 2014; Vogeler et al., 2016).

Repetitive transcranial magnetic stimulation (rTMS) is a promising intervention for smoking cessation. rTMS is a type of non-invasive brain stimulation that uses a time-varying magnetic field to induce electrical currents in the brain. In principle, this can be used to modulate activity within or functional connectivity between regions that are implicated in cigarette smoking. One rTMS protocol using a proprietary deep rTMS coil achieved FDA-clearance in 2021, following a multicenter randomized clinical trial demonstrating that 19.4% of patients remained abstinent at week 18 of the trial in the intent-to-treat analysis (Zangen et al., 2021). Although promising, the smoking cessation rate reported in this trial is comparable to or lower than that produced by varenicline (C. J. Jordan & Xi, 2018) and is significantly lower than response rates following rTMS for other indications such as treatment-resistant depression (e.g., 40-60%; Apostol et al., 2026 and as high as > 80%; Cole et al., 2020, 2022). Further, a replication study failed to find a difference between the active and sham groups (Bellini et al., 2024), implying that individual differences may be an important determinant of the rTMS protocol’s effectiveness.

While the only FDA-cleared protocol relies on a unique, proprietary coil shape, the more commonly used figure-eight coil has also shown considerable promise. Indeed, past work has indicated that rTMS using conventional figure-eight coils can reduce the severity of cigarette craving (Abdelrahman et al., 2021; Amiaz et al., 2009; Apostol et al., 2023; Chang et al., 2018; Flores-Leal et al., 2015; Johann et al., 2003; T. Jordan et al., 2024; X. Li et al., 2013, 2020; Pripfl et al., 2014; Rakesh et al., 2024; Shevorykin et al., 2022; Upton et al., 2023), heaviness of smoking (Abdelrahman et al., 2021; Amiaz et al., 2009; Eichhammer et al., 2003; X. Li et al., 2020; Prikryl et al., 2014; Upton et al., 2023), and withdrawal (Petersen et al., 2025). Furthermore, active rTMS with figure-eight coils has led to higher smoking cessation rates compared to sham rTMS (X. Li et al., 2020; Sheffer et al., 2018).

Despite these documented clinical effects, it is currently unknown which factors predict response to rTMS for smoking cessation. In other words, there may be neurobiological, psychological, sociodemographic, or behavioral factors that predict who experiences reductions in nicotine withdrawal or cigarette craving following rTMS. Because rTMS is a particularly time-intensive treatment, prospectively identifying patients who are likely to respond (or not), rather than relying on trial-and-error, may speed the time to successful smoking cessation and reduce burden on providers. Therefore, there is an urgent need to systematically explore the factors associated with rTMS response, which will help improve smoking cessation outcomes by informing treatment stratification and personalization.

Past literature has identified several behavioral and neurobiological factors that may be associated with rTMS response in substance use disorder populations. Studies of rTMS treatment for stimulant use disorder suggest that rTMS may be more effective in individuals with lower levels of depression and anxiety (Chen et al., 2020; Pettorruso et al., 2019). Studies focused on tobacco use disorder *per se* suggest stronger responses in those who smoke more cigarettes (X. Li et al., 2013) and have greater levels of nicotine dependence (Addicott et al., 2015). Additionally, there may be age-dependent variability in rTMS efficacy (Abdelrahman et al., 2021; Beuzon et al., 2017; Fregni et al., 2006; Kozel et al., 2000; Pallanti et al., 2012).

The neurobiology of tobacco craving, cessation, and relapse has been relatively well-defined, enabling *a priori* hypotheses of neuroimaging-based predictors of rTMS response in the context of smoking cessation. The insula is involved in sensorimotor, social, emotional, attention, and salience processes (Uddin et al., 2017), is a critical brain region in tobacco use. Pioneering work indicated that damage to the insula via stroke in humans led to immediate and lasting smoking cessation, with one patient reporting that his “body forgot the urge to smoke” (Naqvi et al., 2007). Subsequent work has further implicated the insula in nicotine withdrawal symptoms (Abdolahi et al., 2015; Carroll et al., 2015; Fedota et al., 2018; Ghahremani et al., 2021), cigarette cravings (Bi et al., 2017; Moran-Santa Maria et al., 2015; M. T. Sutherland et al., 2013), cigarette consumption (Y. Li et al., 2017; Morales et al., 2014; Stoeckel et al., 2016; Zanchi et al., 2015; Zhou et al., 2017), tobacco cue-reactivity (Janes et al., 2015), and smoking cessation outcomes (Gaznick et al., 2014; Jordan et al., 2024; Joutsa et al., 2022; Suner-Soler et al., 2012). Notably, greater insula functional connectivity and tobacco cue-induced reactivity are associated with successfully quitting smoking (Addicott et al., 2015; Gilman et al., 2018; Janes et al., 2010; Wilcox et al., 2017). Moving beyond the insula, the dorsolateral prefrontal cortex (DLPFC) plays a key role in executive functions and top-down inhibitory control (Jung et al., 2022) and is implicated in cigarette smoking (Franklin et al., 2021). A key finding from previous literature is that reductions in drug craving following rTMS is associated with functional connectivity between the DLPFC and regions implicated in reward valuation, such as the orbital middle prefrontal cortex (OMPFC; X. Li et al., 2017), and regions implicated in attentional processes, such as the inferior parietal lobule (IPL; Su et al., 2020).

To explore the factors associated with rTMS response, we performed a randomized, single-blind, within-subjects, active-control, crossover rTMS experiment. Based on our previous results in a subset of the same research participants (Apostol et al., 2023), we hypothesized that rTMS to the left DLPFC would lead to significant reductions in cigarette craving and rTMS to a control region (left visual cortex; V5) would not. Next, we hypothesized that rTMS responders and nonresponders would differ in terms of depression symptoms, anxiety symptoms, nicotine dependence, number of cigarettes smoked per day, and age. Finally, we hypothesized that responders and nonresponders would differ based functional connectivity from the stimulated region (left DLPFC) to the left OMPFC and left IPL, and display increased functional connectivity from the insula to other brain regions. These hypotheses were pre-registered prior to the completion of data collection.

## Methods

### Participants

Participants (*N* = 61) were adults with TUD (average age: 33.33 +/− 6.66 years; *n* = 31 males and *n* = 29 females) who reported smoking at least five cigarettes per day for 1 year, had positive urinary cotinine measures, fluently spoke English, were right-handed, and were not undergoing smoking cessation treatment. Participants were excluded if they met criteria for other substance use disorders in the past six months, or other psychiatric disorders assessed with the MINI International Neuropsychiatric Interview (MINI; Lecrubier et al., 1997), which assess for major depressive episode/disorder, suicidality/suicide behavior, manic/hypomanic episode, bipolar I/II disorder, other specified bipolar and related disorder, panic disorder, agoraphobia, social anxiety disorder, obsessive compulsive disorder, post-traumatic stress disorder, alcohol use disorder, non-alcohol substance-use disorder, schizophrenia, any other psychotic disorder, major depressive disorder with psychotic features, bipolar I disorder with psychotic features, anorexia nervosa, bulimia nervosa, binge-eating disorder, generalized anxiety disorder or antisocial personality disorder). Participants were also excluded for serious medical illnesses (e.g., neurological, cardiovascular, hepatic, renal, autoimmune diseases, cancer), positive drug tests for other substances besides nicotine and cannabis, pregnancy/breastfeeding, and magnetic resonance imaging (MRI) or rTMS contraindications. One participant was excluded due to missing data, yielding a final sample of *N* = 60 included in the analyses.

### Materials and Supplies

Participants completed questionnaires to assess nicotine dependence and withdrawal, substance use, mood symptoms, demographics, medical history, psychiatric history, education, and sleepiness. Craving, withdrawal, and mood assessments were completed at a baseline in-person screening session, and before and after rTMS treatment sessions. The Shiffman-Jarvik Withdrawal Scale (SJWS) measured nicotine withdrawal and included subscales that measured craving, psychological withdrawal, physical withdrawal, stimulation/sedation, and appetite (Shiffman & Jarvik, 1976). The Urge to Smoke (UTS) scale measured feelings of cigarette craving (Jarvik et al., 2000). The State-Trait Anxiety Inventory (STAI) form Y-1 was collected as a measure of state anxiety (Speilberger et al., 1983). The Fagerstrom Test for Nicotine Dependence (FTND) was a measure of tobacco consumption and dependence (Heatherton et al., 1991). Measures of depression symptoms were collected and included the researcher-administered Patient Health Questionnaire (PHQ; Kroenke et al., 2001) and the Beck Depression Inventory (BDI; Beck et al., 1987). The Positive and Negative Affect Scale (PANAS) measured positive and negative emotions (Watson et al., 1988). The Stanford Sleepiness Scale measured level of arousal and sleepiness (Maclean et al., 1992).

Participants completed a substance use questionnaire that measured whether they had previously used cannabis, cocaine, amphetamines, methamphetamine, hallucinogens, inhalants, antidepressants, sedatives, heroin, methadone, opiates/opioids, or barbiturates. In the case of previous use of a substance, participants were asked how much they used the substance, how often, and whether they were still using it. Participants completed a nicotine-specific questionnaire that measured family nicotine use, the desire to quit, initiation of tobacco use, current use, and withdrawal symptoms. Additionally, a neurological history was completed that assessed past head injuries, hospital visits or medications for emotional problems, neurological diseases, or past treatment for drug addiction. Finally, demographics were collected including biological sex, gender identity, race, ethnicity, and education level. All questionnaires were administered by a member of the research team in a structured clinical interview or completed by participants using a tablet (Apple, Cupertino, CA, USA).

Measures of expired carbon monoxide (CO) and carboxyhemoglobin (COHb) were collected using a Micro+ Smokerlyzer CO monitor (Bedfont Scientific, Harrietsham, England). An Alco-Sensor FST breathalyzer was used to measure recent alcohol use (Intoximeters, St. Louis, MO, USA). Urine samples were collected and assessed for pregnancy (Consult Diagnostics, McKesson Corporation, Irvine, TX, USA), other substances using eight-panel binary lateral flow immunoassay (Alere Toxicology Services, Waltham, MA, USA), and cotinine (Accutest, Jant Pharmacal Corporation, Encino, CA, USA). In the event of a positive urine drug test for cannabis, additional measures were taken to verify that participants were not acutely intoxicated (e.g., Drager DrugTest 5000 saliva tests; Drager-werk, Lubeck, Germany).

### Procedure

This study involved a within-subjects, single-blind, active-controlled, randomized, crossover experiment. Participant recruitment was performed with ads posted to Craigslist, Reddit, Twitter, Facebook, Instagram, and flyers posted to local businesses that sold cigarettes in Los Angeles from April 2019 - July 2024. A structured phone interview was first performed to inform participants about the study procedures and to assess initial eligibility. Then, eligible participants completed an in-person screening appointment that included obtaining informed consent, completing structured clinical interviews, self-report questionnaires, measures of recent drug/alcohol use, tobacco use measures, and MRI scans.

Participants who were eligible to continue in the study after the in-person screening completed two rTMS sessions separated by a minimum of 24 hours to provide a suitable washout (no upper limit). They were allowed to smoke *ad libitum* on the days between appointments but were asked to remain abstinent from cigarettes and other forms of tobacco for at least 12 hours before each rTMS appointment. Abstinence was biochemically verified using a Smokerlyzer and self-report last cigarette time was recorded. If participants were unable to remain abstinent for 12 hours before each rTMS appointment, the visits were rescheduled. At each appointment, participants completed measures of expired CO and COHb, urine drug/pregnancy tests, vitals, questionnaires (SJWS, UTS, STAI, PANAS), MRI scans, and rTMS to the left DLPFC or V5. The questionnaires and MRI scans were performed before and after each rTMS session. The order of stimulation was randomized and counterbalanced using a random number generator. Participants completed a 15-minute rTMS session as described in the below TMS procedures section. Each appointment lasted for 3-4 hours. Participants were paid in cash for their participation in the study and transportation was arranged on a case-by-case basis. All data collection occurred at the Semel Institute for Neuroscience and Human Behavior at University of California, Los Angeles (UCLA).

### Ethics Statement

Prior to data collection, approval was received from the UCLA Institutional Review Board (18-000509-CR-00005). This study was performed in accordance with the Declaration of Helsinki (Association et al., 2013). Written, informed consent was obtained prior to data collection.

### MRI Data Acquisition

MRI scans were performed at the UCLA Staglin Center for Cognitive Neuroscience or the Ahamason-Lovelace Brain Mapping Center. Both scanning centers house a Siemens 3-Tesla Prisma Fit MRI scanner which were outfitted with a 32-channel head coil. At the baseline in-person screening appointment, a T1-weighted structural scan [echo time (TE) = 2.24ms; repetition time (TR) = 2,400ms; voxel resolution = 0.8 × 0.8 × 0.8mm, flip angle=8°] and an 8-minute T2*-weighted multiband resting-state functional image were collected [TE = 37ms; TR = 800ms; field of view = 208mm; slice thickness = 2mm; number of slices = 72; voxel Resolution = 2×2×2mm; acceleration factor = 8, flip angle=52°]. The functional scan was also collected before and after each rTMS session. Imaging data were used for TMS targeting (see TMS Target Localization in Methods) and in the primary analyses (see Statistical Analysis Plan in Methods). MRI scans were performed at the baseline in-person screening, and immediately before and after each rTMS session.

### MRI Data Preprocessing

MRI data were preprocessed using fMRIB Software Library (FSL) v.6.0.4 (Jenkinson et al., 2012). The FSL brain extraction tool (Jenkinson et al., 2005) was used to skull-strip the structural T1-weighted images and FSL’s MCFLIRT tool was used for motion correction of the functional T2*-weighted images (Jenkinson et al., 2002). B0 unwarping and slice-timing correction were performed (Jenkinson et al., 2003). Co-registration of each participant’s functional image to their structural image was performed using FSL linear registration (Andersson et al., 2007). Subsequently, FSL’s FNIRT tool was used to register and normalize the skull-stripped functional and structural images to standard space, which were then smoothed using a 5mm full-width at half-maximum smoothing kernel (Jenkinson et al., 2012). FSL’s MELODIC independent component analysis (ICA) tool was used to deconstruct the functional images into 20 independent components. ICA-FIX was used for denoising of the resting state fMRI scans (Smith et al., 2004).

### TMS Target Localization, Parameters, and Procedures

Personalized DLPFC and V5 stimulation targets were generated for each participant. The DLPFC target within the left DLPFC region of interest (ROI; https://neurosynth.org/analyses/ terms/dlpfc/; retrieved 26 March 2019) was the voxel with maximum functional connectivity to the executive control network that was defined by MELODIC ICA. As an alternative to sham stimulation, active stimulation was applied to the left V5. The V5 stimulation target was the voxel within the left V5 (https://neurosynth.org/analyses/terms/v5/; retrieved 25 March 2019) with maximum functional connectivity to the primary visual network. Details of the personalized targeting method are provided in Petersen et al. (2025). See **Figure 1** for a visualization of the rTMS targets.

**Figure 1.**
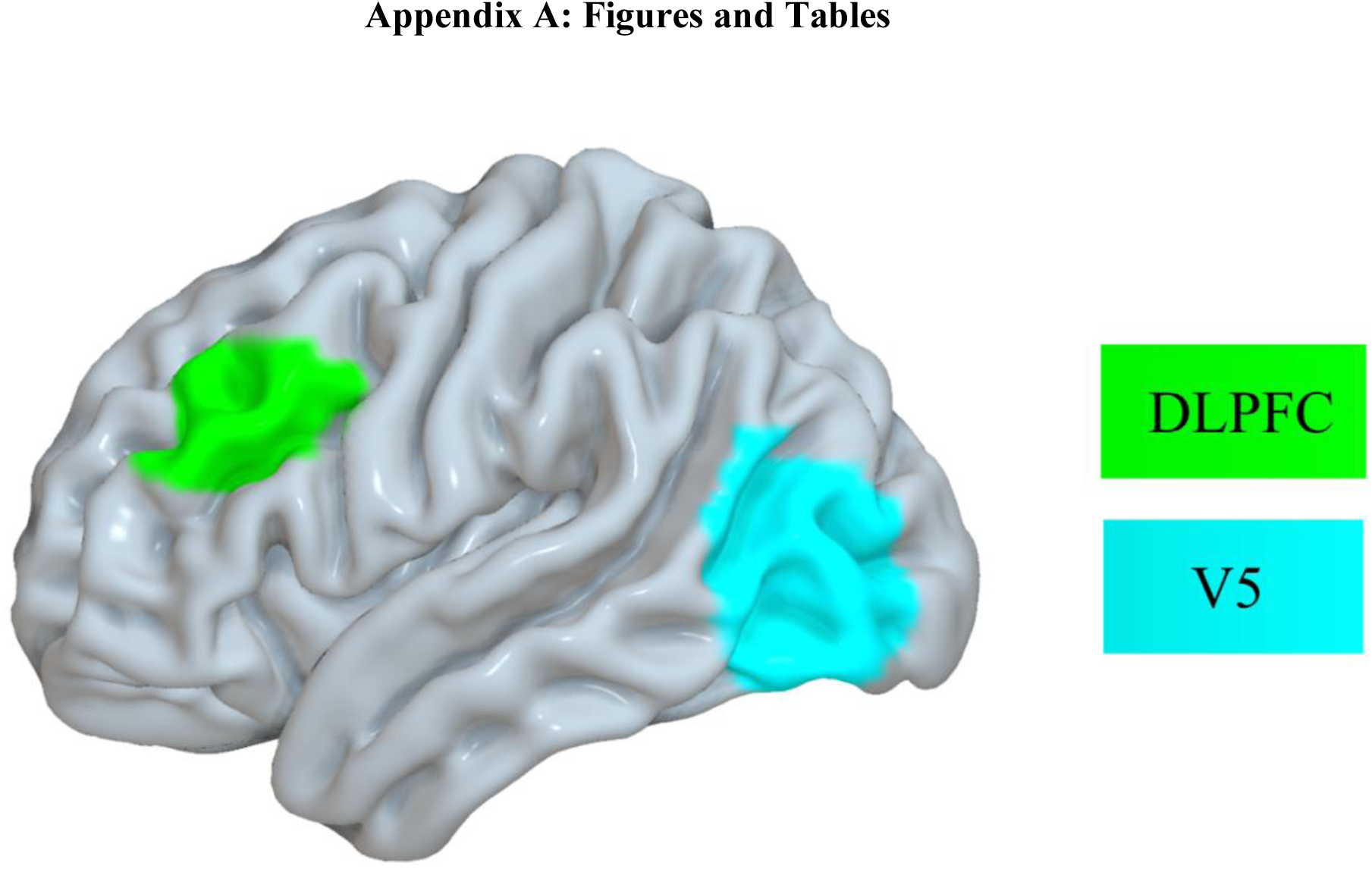
rTMS was applied to the left dorsolateral prefrontal cortex (DLPFC; green) as the experimental stimulation target and the left visual cortex (V5; blue) as the control stimulation target.

rTMS was applied using either a Magstim Super Rapid2 Plus1 stimulator (Roseville, MN, USA) paired with visor2 neuronavigation (ANT Neuro, Hengelo, Netherlands) or a Magventure Magpro X100 stimulator with a Cool-B65 coil (Alpharetta, GA, USA) paired with Brainsight neuronavigation (Rogue Research, Montreal, Quebec, Canada). The rTMS parameters were identical for both devices: 10 Hz, 5 seconds on / 10 seconds off, 60 trains, 3000 pulses total, applied at 100% active motor threshold (aMT). Prior to the first treatment, aMT was calculated by applying single pulses to the left motor cortex (through visual identification of the central sulcus and the precentral gyrus in each participant’s MRI scan). aMT was defined as the stimulation intensity that produced motor evoked potentials in the right abductor pollicis brevis muscle in at least 5 out of 10 trials. At each rTMS session, participants were asked to relax in a chair and remain as still as possible. Neuronavigation was used to position the rTMS coil over each participants individualized DLPFC or V5 targets using an infrared camera that localized the participant’s head in space relative to the TMS coil. Treatments were initiated by a physician at the UCLA TMS Clinical and Research Service, and participants were monitored for tolerability and adverse events. If any signs of adverse events were observed, treatments were stopped and participants underwent assessments by physicians. For each rTMS session, the coil distance-to-scalp, rotation, and angle were collected.

### Statistical Analysis Plan

The hypotheses and analysis plan for this study were pre-registered prior to the completion of data collection and are available at the Open Science Framework data repository (behavioral analyses: https://osf.io/m58rs/ and neuroimaging analyses: https://osf.io/39kvr). Exploratory analyses were also performed and labeled as such. Adjustments from the pre-registration plan are described in the discussion section.

#### Effects of rTMS to DLPFC vs. V5

First, mixed effects linear models were built to confirm that DLPFC stimulation reduced cigarette craving. Notably, a subset of participants included in the present study were also included in previous reports demonstrating reductions in cigarette craving following DLPFC stimulation (Apostol et al., 2023; Petersen et al., 2025). Models were built with rTMS target (DLPFC/V5) and timepoint (pre/post rTMS) as fixed effects, participants as random effects, and the following outcome variables in separate models: SJWS overall scores, SJWS craving subscale, SJWS psychological subscale, SJWS physiological subscale, SJWS stimulation/sedation subscale, and SJWS appetite subscale. Bonferroni correction was applied across these six models (.05 / 6) yielding an adjusted significance threshold of .008. These models were built using the *lmerTest* package in R version 4.4.1 (RStudio Team, 2023). Based on these results, “responders” were defined as participants who experienced any reduction in SJWS craving subscale scores, and “nonresponders” had an increase or no change in SJWS craving subscale scores.

#### Behavioral Data Analysis

Linear models were built using the *lm* command in R to compare behavioral variables that were hypothesized to differ significantly between responders and nonresponders. Models were built with the following outcome variables: PHQ, BDI, STAI, FTND, number of cigarettes in the past 30 days, and age. Additional behavioral variables were collected in this study besides the variables included in the hypotheses. For these variables, rather than computing null hypothesis significance tests with a risk of inflating Type 1 Error, two separate approaches were taken for continuous versus categorical variables. For continuous variables, 95% confidence intervals were computed using the *rstatix* (Kassambara, 2020) and *gtsummary* (Sjoberg et al., 2021) packages in R, then effect sizes (Cohen’s d) were computed using the *cohens_d* command. For categorical variables, Fisher’s exact test, chi square tests, and effect sizes (Cramer’s V) were computed using the *fisher.test*, *chisq.test*, and *cramer_v* commands in R, respectively.

#### Neuroimaging Data Analysis

Three participants were excluded from the neuroimaging analyses due to incomplete fMRI scans. Pre-registered region-of-interest (ROI)-ROI analyses were performed to compare functional connectivity between the stimulated region (DLPFC) and brain regions hypothesized to be associated with reductions in drug craving following rTMS (OMPFC and IPL). The DLPFC mask was generated from Neurosynth (https://neurosynth.org/analyses/terms/dlpfc/), thresholded to z = 5, lateralized to the left hemisphere, and binarized using *fslmaths*. The OMPFC mask was created using *fslmaths* and was a 5mm sphere centered at MNI (−39, 45, −9) in line with (X. Li et al., 2017) and was binarized. The IPL mask was created using *fslmaths* and consisted of a 5mm sphere centered at MNI (−52, −58, 46) in line with (Su et al., 2020) that was binarized. Once the masks were created, mean ROI timeseries were extracted using *fslmeants*.

Correlations were calculated between the time-series and then z-transformed, then *t*-tests were performed between responders and nonresponders using R. Functional connectivity between the DLPFC-OMPFC and DLPFC-IPL was compared between responders and nonresponders at two timepoints. Pre-rTMS functional connectivity was first assessed between groups, then the change in functional connectivity from pre-rTMS to post-rTMS was compared between responders and nonresponders.

Pre-rTMS insula-whole brain functional connectivity patterns were then compared between responders and nonresponders. Based on a probabilistic insula atlas from Faillenot et al. (2017), 14 bilateral insula sub-region masks were constructed: anterior long gyrus, anterior inferior cortex, posterior short gyrus, middle short gyrus, anterior short gyrus, and posterior long gyrus. Whole left and right insula masks were created by combining these sub-regions using *fslmaths*. The 14 insula ROIs were transformed to 2mm MNI space, thresholded at 25%, and binarized.

Each subject’s preprocessed functional scans were then normalized to MNI space using *applywarp*, insula time series were extracted for each of the 14 insula masks using *fslmeants*, then seed-based z-maps were created using *fsl_glm*. The z-maps were merged into a 4D file using *fslmerge*, and these merged z-map files were used in the *randomise* command (Winkler et al., 2014). The design matrix and contrast files were built to test two contrasts: responder > nonresponder and nonresponder > responder. *Randomise* was run separately for each of the 14 insula ROIs with 5000 permutations per run. Threshold-free cluster enhancement (TFCE) correction was applied for each *randomise* analysis.

#### Exploratory Analyses

To supplement the pre-registered analyses, additional exploratory analyses were performed. First, the *randomise* pipeline was run with a left DLPFC seed to explore potential connectivity differences between responders and nonresponders from the stimulated brain region. Next, given the arbitrary nature of the responder/nonresponder delineation (responders = any reduction in craving; nonresponders = increase or no change in craving), the *randomise* analysis pipeline was run with a new criteria for “responders” and “nonresponders” such that responders (*n* = 20) were defined as participants who had a change of at least one point in the SJWS craving subscale (maximum SJWS craving subscale score reduction = 2.8; minimum reduction = 0.2) and nonresponders had less than a one point change in the SJWS craving subscale (*n* = 37). In other words, the “responders” were defined as the ∼1/3 of participants who experienced the greatest reduction in cigarette craving for this exploratory analysis. These analyses were run with the left and right whole insula seed regions. Finally, ROI-ROI functional connectivity was computed between the left and right whole insula and the following regions that are key components of addiction circuitry: nucleus accumbens (NAc; Balfour, 2004), anterior cingulate cortex (ACC; Ghahremani et al., 2021), and hippocampus (Kutlu & Gould, 2016) defined by the Hammers atlas (Hammers et al., 2003), in addition to the stimulated region (left DLPFC) using the DLPFC ROI from Neurosynth (https://neurosynth.org/analyses/ terms/dlpfc/; retrieved 26 March 2019). Functional connectivity differences between responders and nonresponders were assessed using *t*-tests. If significant differences were identified between responders and nonresponders for any of the above ROIs, then functional connectivity for that ROI was computed with the 12 insula sub-regions, including the left/right hemisphere anterior long gyrus, anterior inferior cortex, posterior short gyrus, middle short gyrus, anterior short gyrus, and posterior long gyrus of the insula. For these exploratory ROI-ROI functional connectivity analyses, Bonferroni correction was applied within each hemisphere.

## Results

### Effects of rTMS to DLPFC vs. V5

Mixed-effects linear models were computed to compare changes in SJWS scores from before to after rTMS to the DLPFC versus V5. Six models were built and the Bonferroni correction was applied (.05 / 6 = .008). With the corrected significance threshold, the SJWS craving subscale model had significant pre/post [*t*(168.235) = −3.239, *p* = 0.001] and TMS target [*t*(169.068) = −2.855, *p* = 0.005], but the interaction term was not statistically significant [*t*(168.235) = 1.254, *p* = 0.211]. Following multiple comparison correction, there were no statistically significant effects for the SJWS psychological, physiological, stimulation/sedation, appetite, or overall models (all *p*-values > .05). See **Table 1** for a summary of the mixed-effects linear models. Post-hoc *t*-tests revealed that stimulation to the DLPFC led to a significant reduction in SJWS Craving scores (*t*(59) = 3.6410, *p* = .0006). Stimulation of V5 did not (*t*(55) = 1.8357, *p* = 0.0718; **Figure 2**). Although the pre-rTMS SJWS Craving scores were numerically higher for the DLPFC session compared to the V5 session, a paired samples *t*-test indicated that they did not differ significantly, *t*(55) = 1.090, *p* = .280. The SJWS craving subscale was the variable that was used to define the “responder” and “nonresponder” groups. “Responders” were defined as participants with a reduction in SJWS craving subscale scores (*n* = 38) and “nonresponders” had worse or no change in SJWS craving subscale scores (*n* = 22; **Figure 3**).

**Figure 2.**
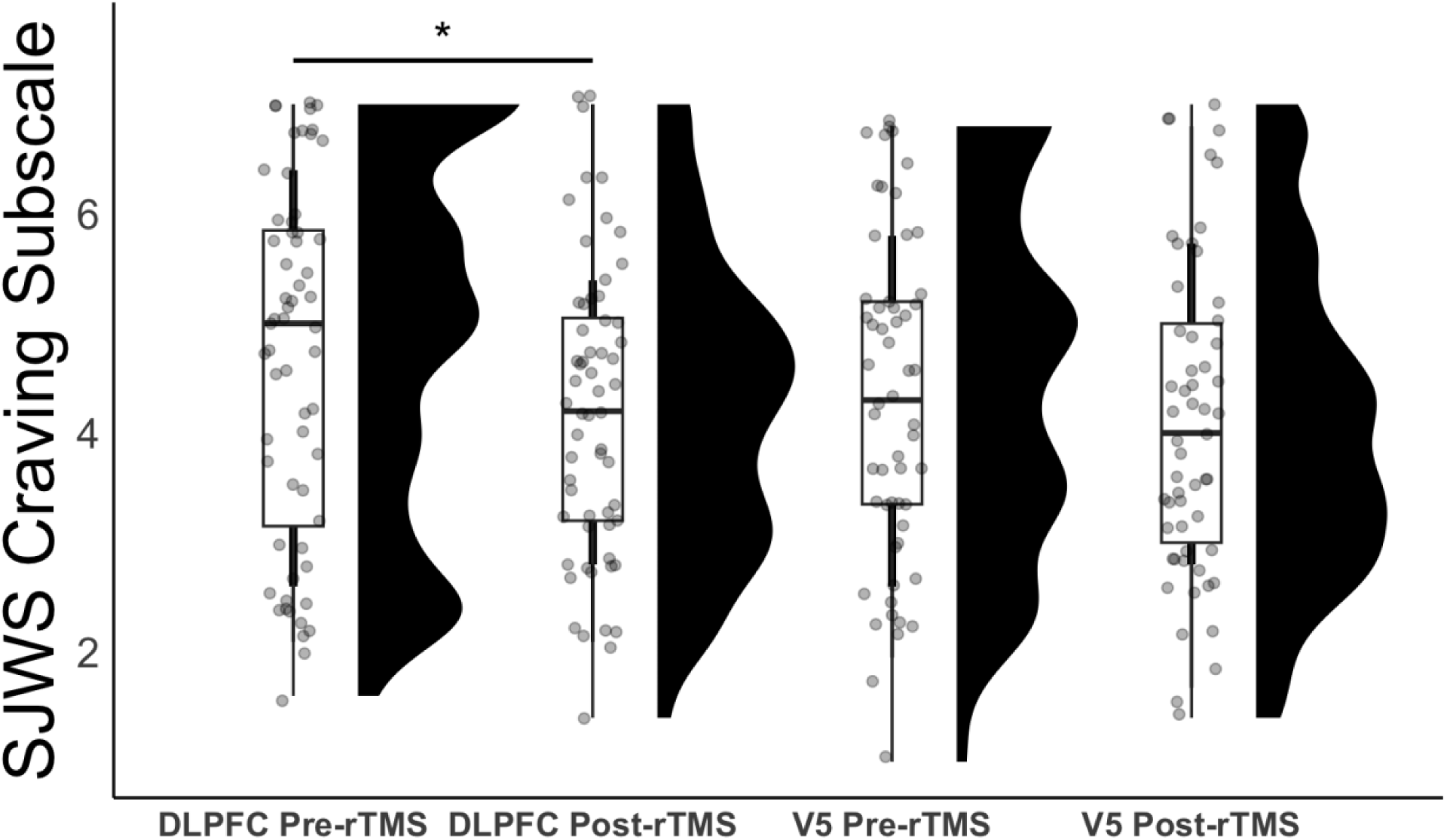
rTMS to the DLPFC led to significant reductions in SJWS craving subscale scores, *p* < .05 (left two box-and-whisker plots). rTMS to V5 did not lead to significant reductions in SJWS craving subscale scores, *p* > .05 (right two box-and-whisker plots). *N* = 60 participants were included in this analysis, which involved building a mixed-effects linear model to compare pre/post rTMS changes in SJWS craving subscale scores following DLPFC vs. V5 stimulation. Mean values (black dots) are displayed in the box-and-whisker plots, which are superimposed over the data points, and the distributions of the data are shown to the right of each box-and-whisker plot. Box-and-whisker plots show the median values, upper and lower quartiles, and the minimum/maximum values. *P < 0.05.

**Figure 3.**
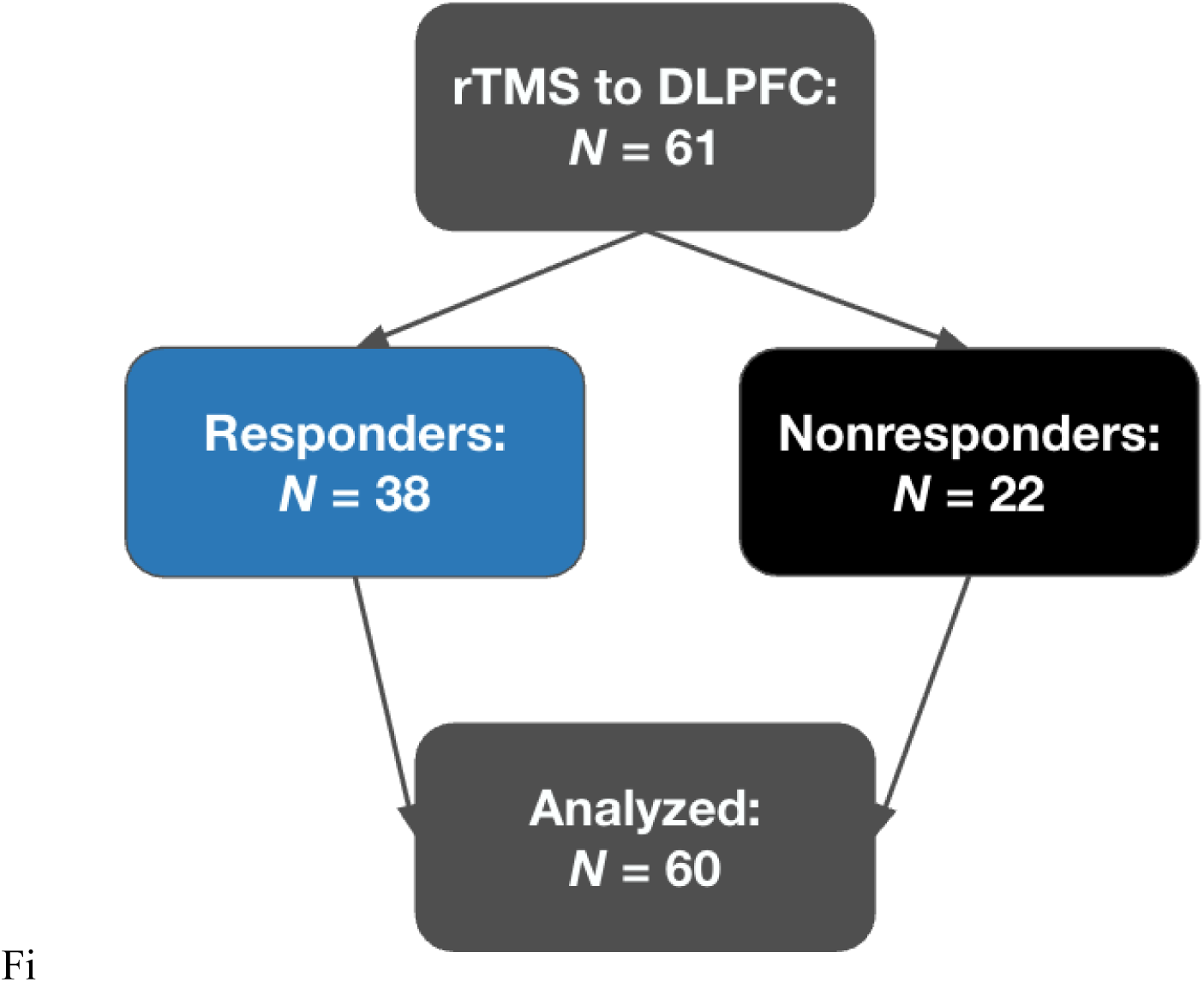
Of the *N* = 61 who were randomized to receive rTMS to the DLPFC, *N* = 60 completed all assessments and were included in the final analysis. “Responders” were defined as participants who experienced a reduction in craving following rTMS to DLPFC (*n* = 38) and “nonresponders” were participants who experienced to change or an increase in craving (*n* = 22). One participant was excluded due to missing data.

**Table 1.** Results from mixed-effects linear models that were built to compare pre/post rTMS changes between DLPFC vs. V5 stimulation with the following outcome variables: SJWS craving subscale, SJWS psychological subscale, SJWS physiological subscale, SJWS stimulation/sedation subscale, SJWS appetite subscale, and SJWS overall scores. rTMS to DLPFC led to significant reductions in SJWS craving subscale scores, whereas rTMS to V5 did not. No other significant effects were observed following Bonferroni correction (.05 / 6).

| Model | Fixed Effect | Estimate | Standard Error | t-value | df | p-value |
| --- | --- | --- | --- | --- | --- | --- |
| SJWS Craving | Pre/Post | -0.460 | 0.142 | -3.239 | 168.235 | 0.001 |
|  | TMS Target | -0.416 | 0.146 | -2.855 | 169.068 | 0.005 |
|  | Interaction | 0.256 | 0.204 | 1.254 | 168.235 | 0.211 |
| SJWS Psychological | Pre/Post | -0.197 | 0.143 | -1.373 | 167.921 | 0.172 |
|  | TMS Target | -0.272 | 0.147 | -1.856 | 169.480 | 0.065 |
|  | Interaction | 0.172 | 0.206 | 0.833 | 167.921 | 0.406 |
| SJWS Physiological | Pre/Post | -0.127 | 0.106 | -1.198 | 169.120 | 0.232 |
|  | TMS Target | -0.153 | 0.109 | -1.402 | 169.890 | 0.163 |
|  | Interaction | 0.247 | 0.153 | 1.613 | 169.120 | 0.109 |
| SJWS Stimulation/Sedation | Pre/Post | 0.250 | 0.251 | 0.995 | 169.043 | 0.321 |
|  | TMS Target | -0.083 | 0.257 | -0.322 | 170.795 | 0.748 |
|  | Interaction | -0.071 | 0.362 | -0.197 | 169.043 | 0.844 |
| SJWS Appetite | Pre/Post | 0.000 | 0.250 | 0.000 | 169.195 | 1.000 |
|  | TMS Target | -0.253 | 0.256 | -0.990 | 170.904 | 0.324 |
|  | Interaction | 0.411 | 0.360 | 1.141 | 169.195 | 0.255 |
| SJWS Overall | Pre/Post | -0.227 | 0.090 | -2.515 | 168.270 | 0.013 |
|  | TMS Target | -0.280 | 0.093 | -3.020 | 169.056 | 0.003 |
|  | Interaction | 0.215 | 0.130 | 1.650 | 168.270 | 0.101 |

### Behavioral Responder/Nonresponder Results

The following *a priori*-defined behavioral variables were compared between responders and nonresponders using linear models: cigarettes per day in the last 30 days, age, PHQ, BDI, STAI, and FTND scores. The number of cigarettes per day in the last 30 days differed significantly between the two groups, *t*(53) = −2.363, *p* = 0.022, such that responders used more cigarettes per day (*M* = 11.44, *SD* = 5.909) compared to nonresponders (*M* = 7.952, *SD* = 4.165). The remaining variables did not differ significantly between responders and nonresponders. Age: *t*(58) = −0.898, *p* = 0.373 (Responders: *M* = 33.921, *SD* = 6.582; Nonresponders: *M* = 32.32, *SD* = 6.813). FTND: *t*(58) = −0.21, *p* = 0.834 (Responders: *M* = 3.974, *SD* = 2.060; Nonresponders: *M* = 3.864, *SD* = 1.754). STAI: *t*(55) = −0.894, *p* = 0.375 (Responders: *M* = 32.389, *SD* =8.706; Nonresponders: *M* = 30.476, *SD* = 5.853). BDI: *t*(56) = −1.12, *p* = 0.268 (Responders: *M* = 5.027, *SD* = 5.449; Nonresponders: *M* = 3.571, *SD* = 3.155). PHQ: *t*(56) = −0.287, *p* = 0.775 (Responders: *M* = 3.108, *SD* = 4.115; Nonresponders: *M* = 2.809, *SD* = 3.188). See **Figure 4** for raincloud plots of these models.

**Figure 4.**
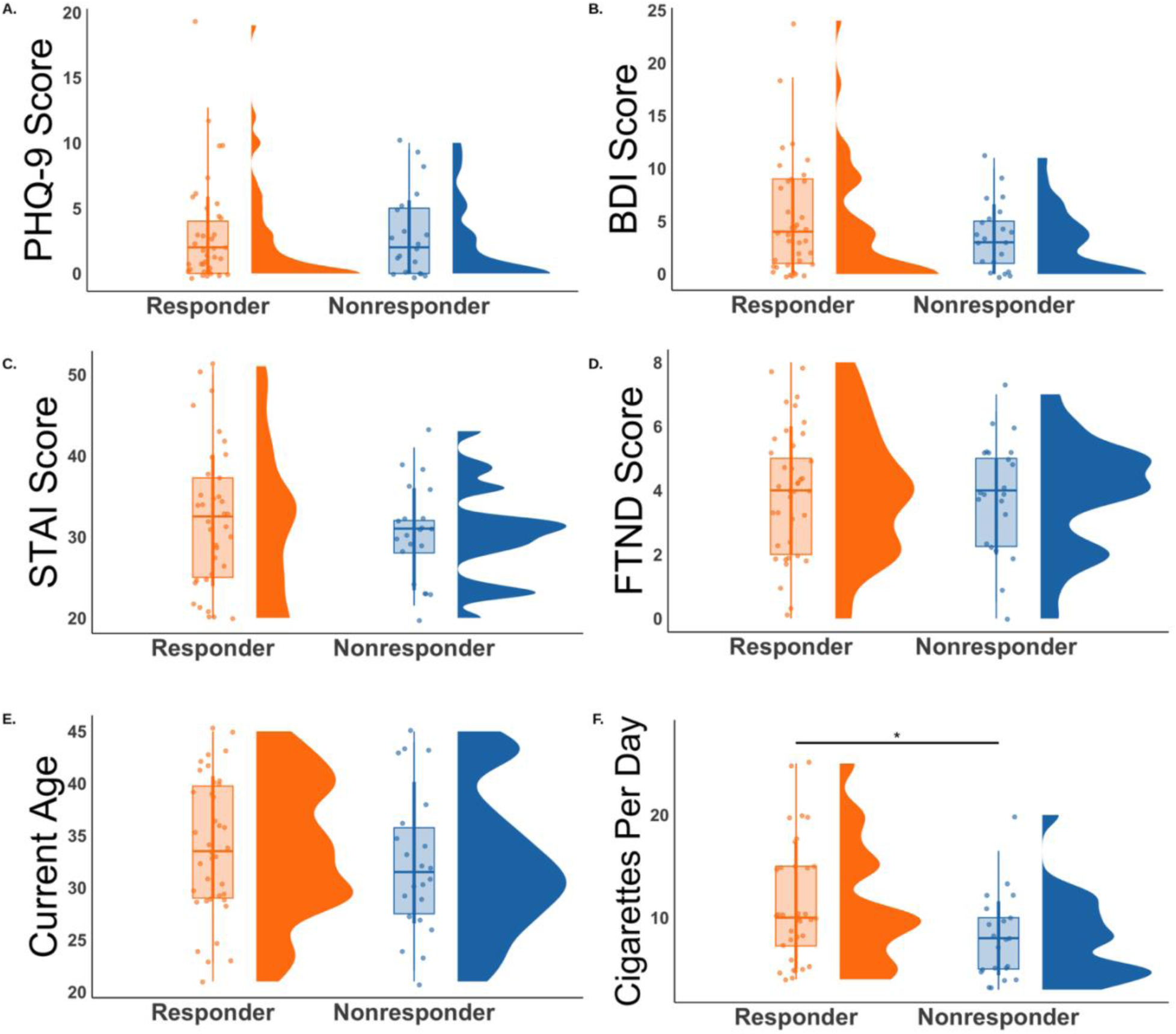
Linear models were built to compare responders and nonresponders based on depression symptoms (PHQ in panel A and BDI in panel B), anxiety symptoms (STAI; panel C), nicotine dependence (FTND; panel D), age (panel E) and number of cigarettes per day (panel F). Responders are shown in orange and nonresponders are shown in blue. Mean values (orange dots for responders; blue dots for nonresponders) are displayed in the box-and-whisker plots, which are superimposed over the data points, and the distributions of the data are shown to the right of each box-and-whisker plot. Box-and-whisker plots show the median values, upper and lower quartiles, and the minimum/maximum values. \**p* < 0.05.

For other behavioral variables collected in this study without *a priori* hypotheses, confidence intervals and Cohen’s d were computed for continuous variables, and Fisher’s exact test and Cramer’s V were computed for categorical variables. Three continuous variables were observed to have large effect sizes between responders and nonresponders immediately prior to rTMS. Baseline SJWS craving subscale scores: responder *M* = 5.242 (95% CI: 4.805, 5.679), nonresponder *M* = 3.664 (95% CI: 2.94, 4.388), Cohen’s *d* = 1.059. Baseline SJWS overall scores: responder *M* = 3.919 (95% CI: 3.625, 4.213), nonresponder *M* = 3.121 (95% CI: 2.64, 3.602), Cohen’s *d* = .803. Baseline UTS scores: responder *M* = 5.066 (95% CI: 4.509, 5.623), nonresponder *M* = 3.359, (95% CI: 2.536, 4.182), Cohen’s *d* = .960. See **Figure 5** for a visualization of these 3 variables with large effect sizes. The remaining variables had small-to-moderate effect sizes. See **Table 2** for the statistics for the continuous variables and **Table 3** for the categorical variables.

**Figure 5.**
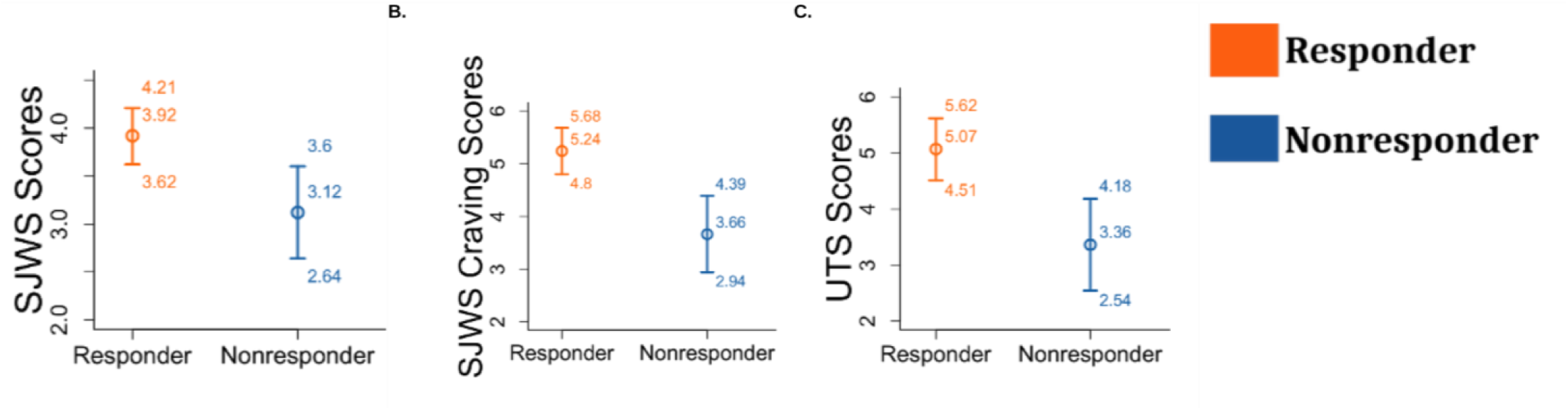
Confidence intervals and effect sizes are displayed for pre-rTMS SJWS overall scores (panel A), SJWS craving subscale scores (panel B), and UTS scores (panel C). These 3 variables were the pre-rTMS measurements. The effect sizes (Cohen’s d) were large for these three variables. Responders are displayed in orange and nonresponders are displayed in blue.

**Table 2.**
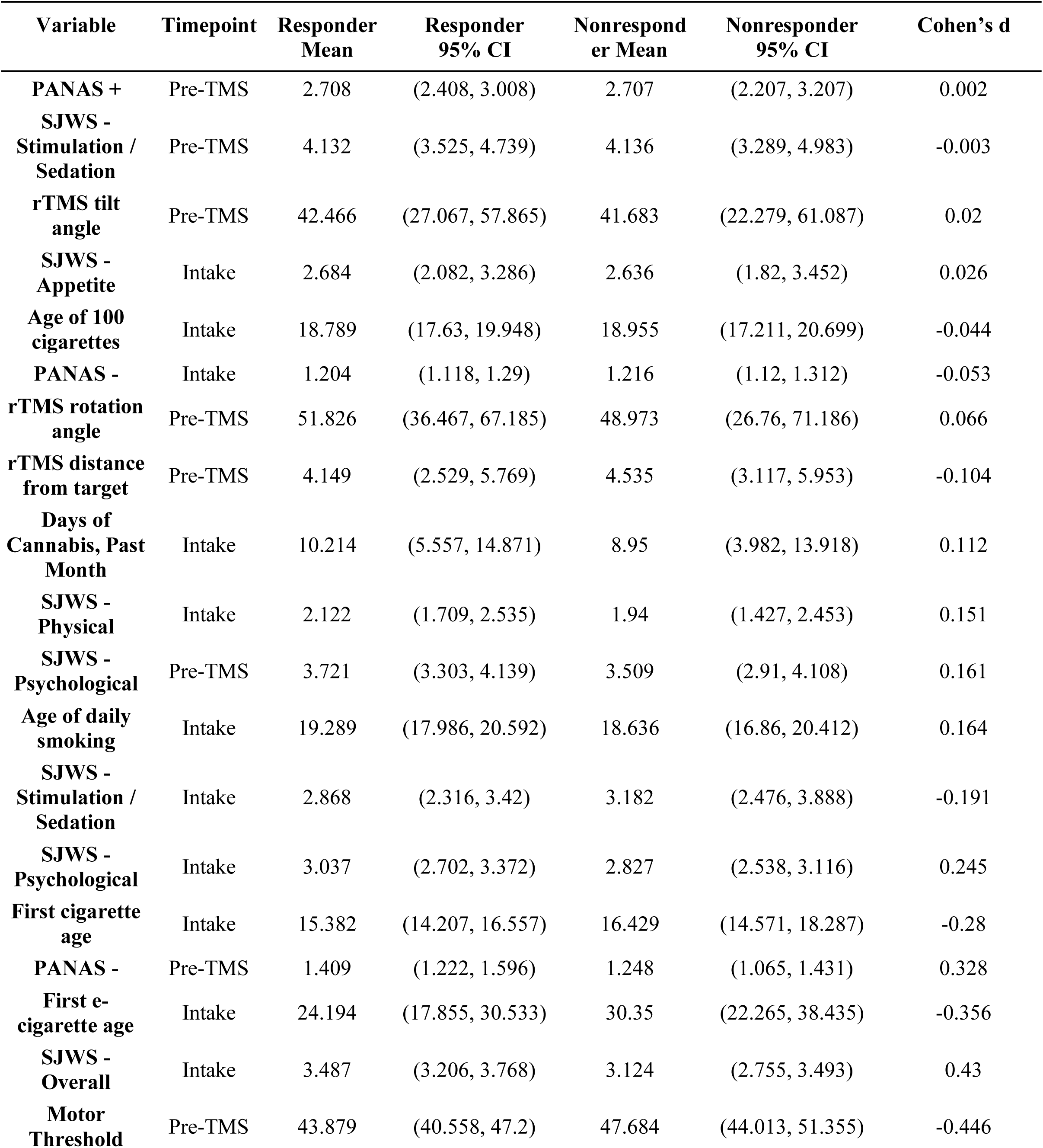
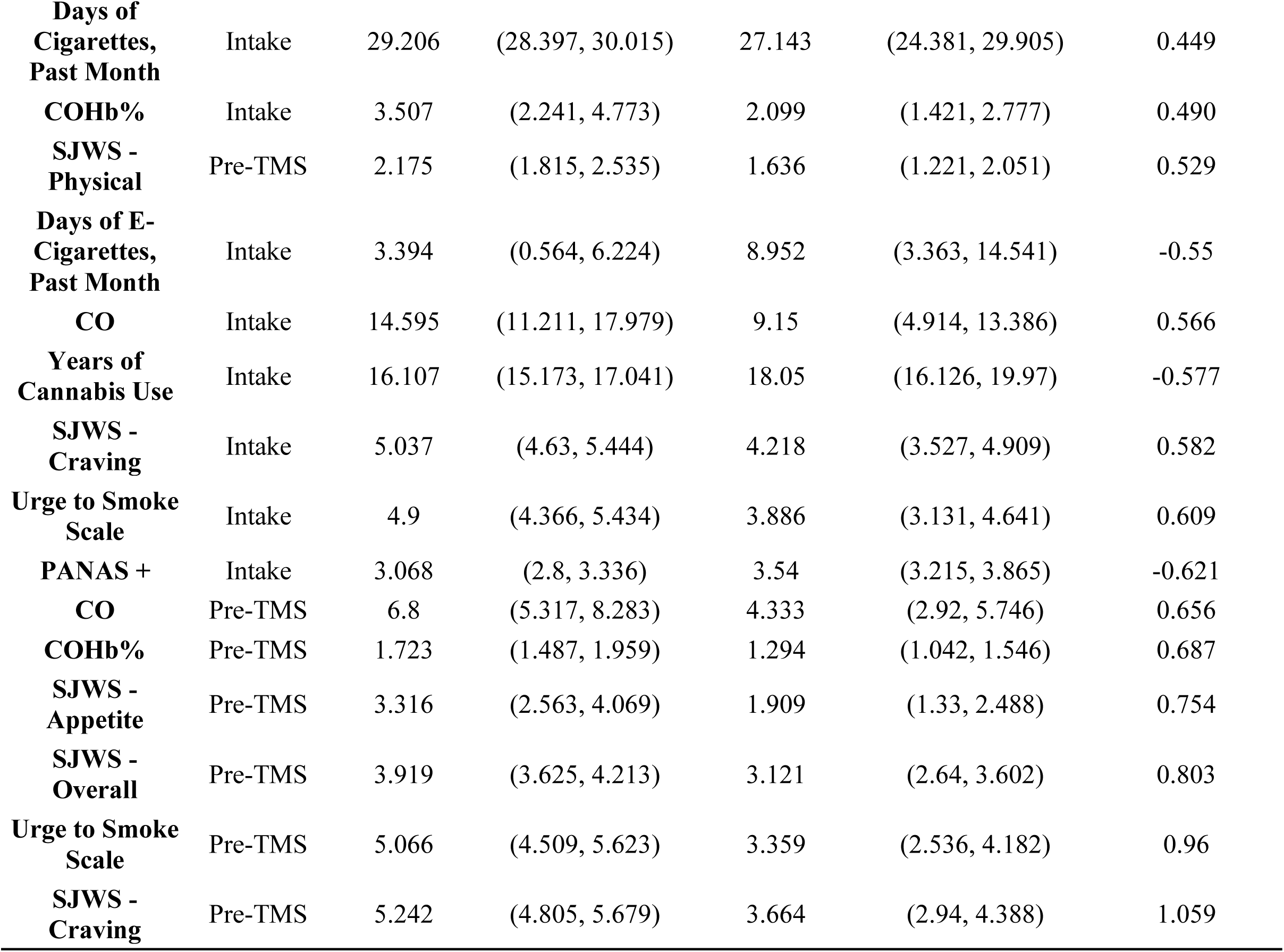
Means, 95% confidence intervals (CIs), and Cohen’s *d* (effect size) for responders and nonresponders. Variable timepoint includes the baseline appointment when participants were smoking ad libitum (“Intake”) or prior to applying rTMS to L-DLPFC following a period of abstinence from smoking (“Pre-TMS”).

**Table 3.** *P*-values from Fisher’s Exact Test and Cramer V values shown for responders and nonresponders. Variable timepoint includes the baseline appointment when participants were smoking ad libitum (“Intake”) or prior to applying rTMS to L-DLPFC following a period of abstinence from smoking (“Pre-TMS”). When cells contain N/A, it indicates homogeneity in responses (e.g, there was no barbiturate use in the sample).

| Variable | Timepoint | Fisher Exact Test *p*-value | Cramer V |
| --- | --- | --- | --- |
| Past cannabis use (yes/no) | Intake | 1 | 0 |
| Past meth use (yes/no) | Intake | 1 | 0 |
| If you quit previously did you experience withdrawal? | Intake | 1 | 0 |
| Past treatment for alcohol or drug abuse | Intake | 1 | 0 |
| Past problem due to abuse of drugs or medications | Intake | 1 | 0 |
| Biological Sex | Intake | 1 | 0 |
| Past head injury involving unconsciousness | Intake | 1 | 0.007 |
| Past cocaine use (yes/no) | Intake | 0.786 | 0.009 |
| Did your Mother smoke? | Intake | 0.783 | 0.015 |
| Do you only smoke cigarettes? | Intake | 0.712 | 0.019 |
| Did your Father smoke? | Intake | 0.787 | 0.023 |
| MINI Diagnoses | Intake | 0.782 | 0.025 |
| Past heroin use (yes/no) | Intake | 0.375 | 0.035 |
| Past methadone use (yes/no) | Intake | 0.375 | 0.035 |
| Did you take cessation medications? | Intake | 0.659 | 0.035 |
| Medications for mental or emotional problems | Intake | 0.698 | 0.041 |
| Past amphetamine use (yes/no) | Intake | 0.58 | 0.065 |
| Hospital visits for mental or emotional problems | Intake | 0.547 | 0.067 |
| Past inhalants use (yes/no) | Intake | 0.459 | 0.088 |
| Past antidepressant use (yes/no) | Intake | 0.459 | 0.088 |
| Depth of Inhalation | Intake | 1 | 0.102 |
| Gender Identity | Intake | 0.869 | 0.106 |
| Would you like to quit? | Intake | 0.367 | 0.107 |
| # of quit attempts | Intake | 1 | 0.114 |
| Past hallucinogens use (yes/no) | Intake | 0.280 | 0.12 |
| Past opiate/opioid use (yes/no) | Intake | 0.170 | 0.159 |
| Ethnicity | Intake | 0.149 | 0.17 |
| Education level | Intake | 0.825 | 0.205 |

**Table 3.** *P*-values from Fisher’s Exact Test and Cramer V values shown for responders and nonresponders.
|  |  |  |  |
| --- | --- | --- | --- |
| Past sedative use (yes/no) | Intake | 0.039 | 0.257 |
| Stanford Sleepiness Scale | Pre-TMS | 0.384 | 0.302 |
| Stanford Sleepiness Scale | Intake | 0.280 | 0.313 |
| People in household who smoke | Intake | 0.044 | 0.427 |
| Race | Intake | 0.026 | 0.432 |
| Past barbiturate use (yes/no) | Intake | N/A | N/A |
| Did you attend cessation programs? | Intake | N/A | N/A |

### Neuroimaging Results

Pre-rTMS functional connectivity between 1) L-DLPFC and L-OMPFC and 2) L-DLPFC and L-IPL was compared between responders and nonresponders. There were no significant differences in pre-rTMS functional connectivity between the groups: L-DLPFC and L-OMPFC pre-rTMS functional connectivity [*t*(48.413) = −1.303, *p* = 0.199]; L-DLPFC and L-IPL pre-rTMS functional connectivity [*t*(42.004,) = 0.923, *p* = 0.361]. The change in functional connectivity from before to after rTMS was then compared between responders and nonresponders, but did not differ significantly between the two groups: L-DLPFC and L-OMPFC functional connectivity change [*t*(39.878) = −0.841, *p* = 0.405]; L-DLPFC and L-IPL functional connectivity change [*t*(38.27) = 0.029, *p* = 0.977]. **Figure 6** displays the results for the DLPFC-OMPFC and DLPFC-IPL analyses.

**Figure 6.**
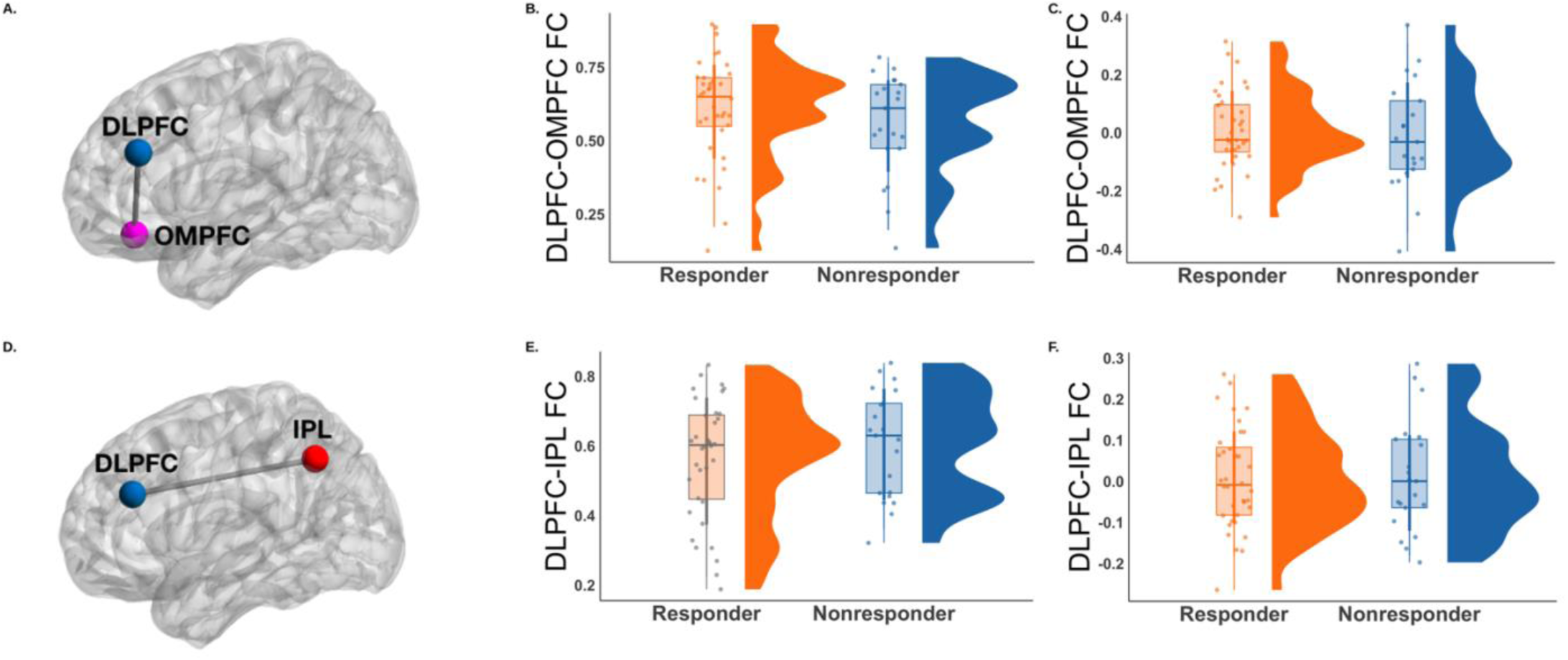
Functional connectivity (FC) was calculated between the DLPFC/OMPFC (panel A) prior to rTMS (panel B), and the pre/post change in functional connectivity (panel C) was computed. Similarly, functional connectivity was calculated between the DLPFC/IPL (panel D) prior to rTMS (panel E), and the pre/post change in functional connectivity (panel F) was computed. There were no significant differences in functional connectivity between the responders (orange) and nonresponders (blue).

Insula seed-based functional connectivity was compared between responders and nonresponders for the following 14 left/right hemisphere regions: whole insula, anterior long gyrus, anterior inferior cortex, posterior short gyrus, middle short gyrus, anterior short gyrus, and posterior long gyrus. No significant effects were observed following TFCE correction for multiple comparisons (all corrected *p*-values > .05) for the responder > nonresponder contrasts. Similarly, no significant effects were observed following TFCE correction for multiple comparisons (all corrected *p*-values > .05) for the nonresponder > responder contrasts.

### Exploratory Results

The *randomise* pipeline was run with a left DLPFC seed to compare left DLPFC connectivity patterns between responders and nonresponders. The analysis revealed that there were no significant differences in left DLPFC seed-based functional connectivity for either the responder > nonresponder contrast (all corrected *p*-values > .05) or the nonresponder > responder contrast (all corrected *p*-values > .05).

Next, left and right insula functional connectivity was compared between responders and nonresponders with “response” redefined as ≥ 1-point reduction in the SJWS craving subscale (rather than as ≥ 0-point reduction in craving in the pre-registered analyses). There were no significant differences between the two groups following TFCE correction for the responder > nonresponder contrast for the left and right whole insula models (all corrected *p*-values > .05). However, the left insula model indicated that there were significant clusters in which nonresponders had greater functional connectivity compared to responders. Two clusters were identified, consisting of 691 (TFCE corrected *p*-value = 0.037; peak MNI coordinate: −2, −98, 6) and 232 (TFCE corrected *p*-value = 0.035; peak MNI coordinate: 4, −62, 64) voxels, which corresponded to the occipital pole and the precuneus, respectively. Four additional clusters of 7, 5, 2, and 1 voxels were identified but not interpreted due to limited spatial extent. Similarly, the right insula model indicated that there were significant clusters in which nonresponders displayed greater functional connectivity compared to responders, including clusters of 396 (TFCE corrected *p*-value = 0.04; peak MNI coordinate: −4, −90, −4) and 187 (TFCE corrected *p*-value = 0.039; peak MNI coordinate: −2, −98, 6) voxels which corresponded to two regions within the occipital pole. One additional cluster of 1 voxel was identified but cannot be interpreted due to limited spatial extent. See **Figure 7** for the left and right insula functional connectivity maps.

**Figure 7.**
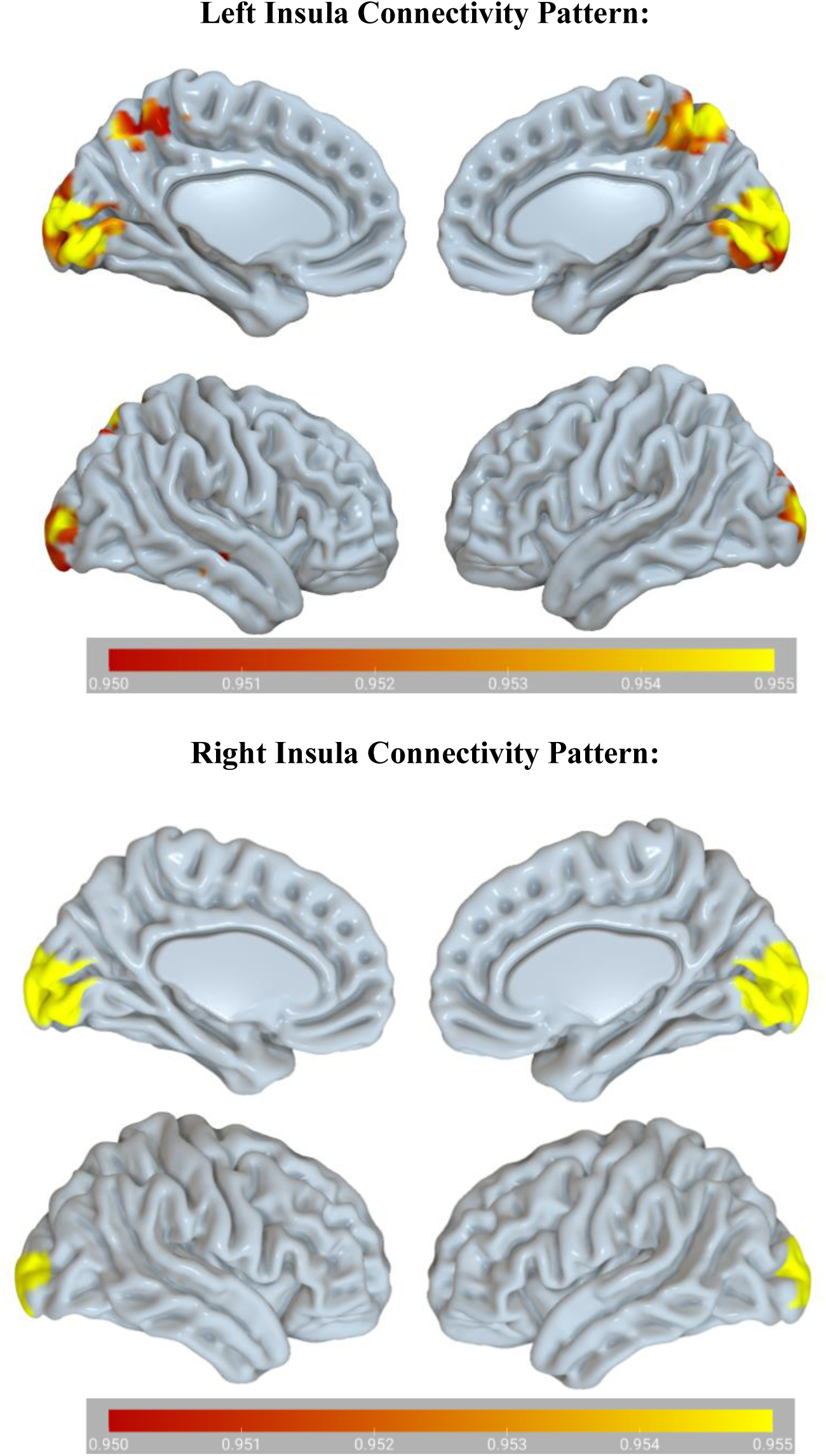
Exploratory insula analyses revealed that nonresponders had greater connectivity between the left insula and occipital and parietal regions (images) compared to responders. Similarly, nonresponders had greater connectivity between the right insula and occipital regions (bottom) compared to responders. The color bars show a range of *p*-values ranging from < .05 to < .001 (1 – *p*).

ROI-ROI analyses were then performed between the left and right whole insula and the following ROIs that are critical for addiction circuitry: NAc, hippocampus, and ACC, in addition to the rTMS stimulation target (left DLPFC). Functional connectivity between the left insula and the right NAc was significantly lower in responders compared to nonresponders, *t*(49.978) = - 3.58, corrected *p*-value = 0.005; Cohen’s *d* = −0.924, 95% CI = −1.500 −0.347. Similarly, functional connectivity between the right insula and right NAc was significantly lower in responders compared to nonresponders *t*(50.577) = −3.204, corrected *p*-value = 0.016; Cohen’s *d* = −0.822, 95% CI = −1.394 - 0.251 (**Figure 8**). Functional connectivity between the left and right insula and the left DLPFC, bilateral hippocampus, and bilateral ACC did not differ significantly between the groups (all *p*-values > .05; see **Table 4**).

**Figure 8.**
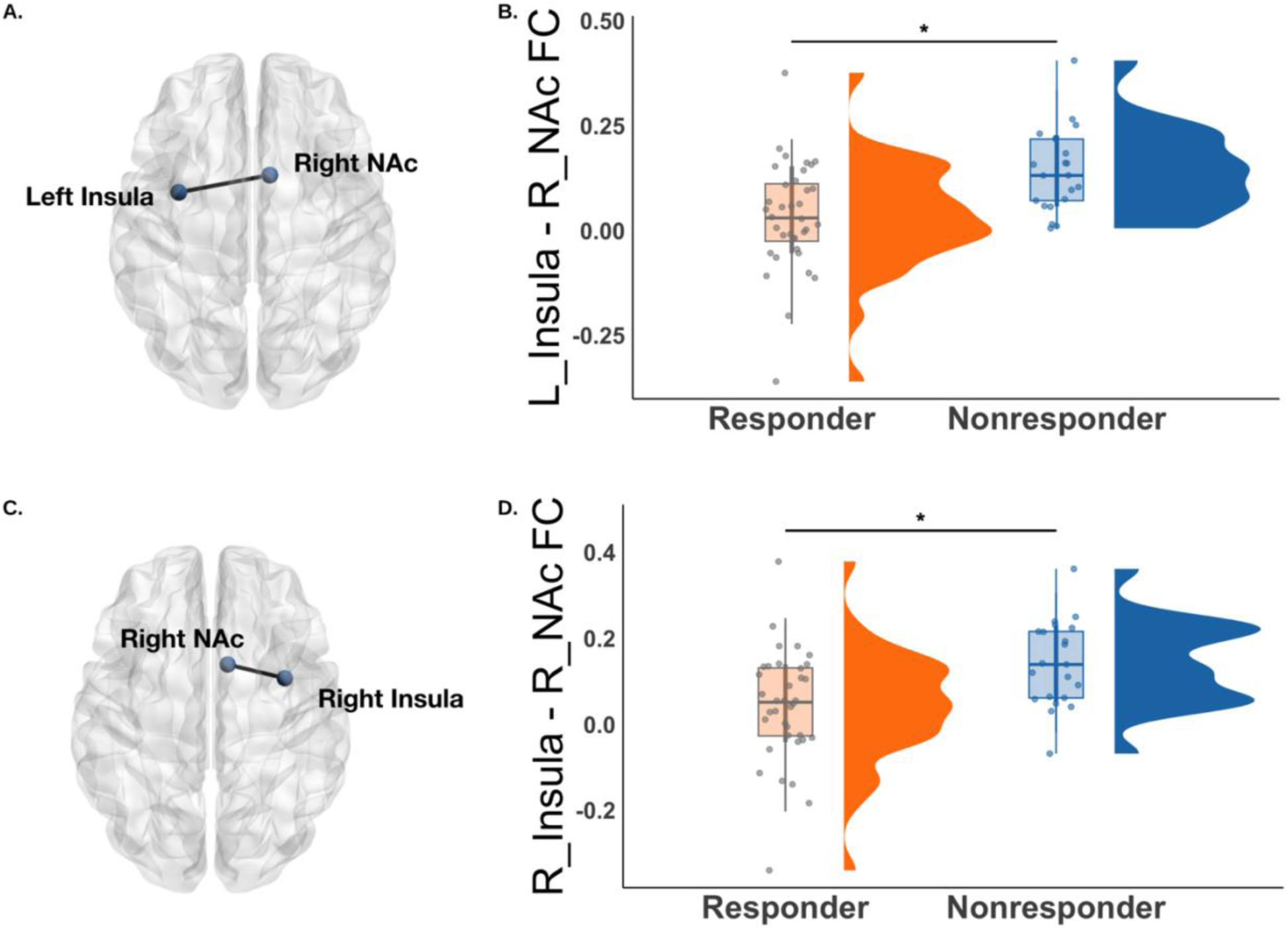
Functional connectivity (FC) between the left insula (L_Insula) and right NAc R_NAc) shown in panel A was significantly lower in responders compared to nonresponders (panel B). Similarly, functional connectivity between the R_Insula and R_NAc (panel C) was significantly lower in responders compared to nonresponders (panel D). Responders are shown in orange and nonresponders are shown in blue. \**p* < 0.05.

**Table 4.**
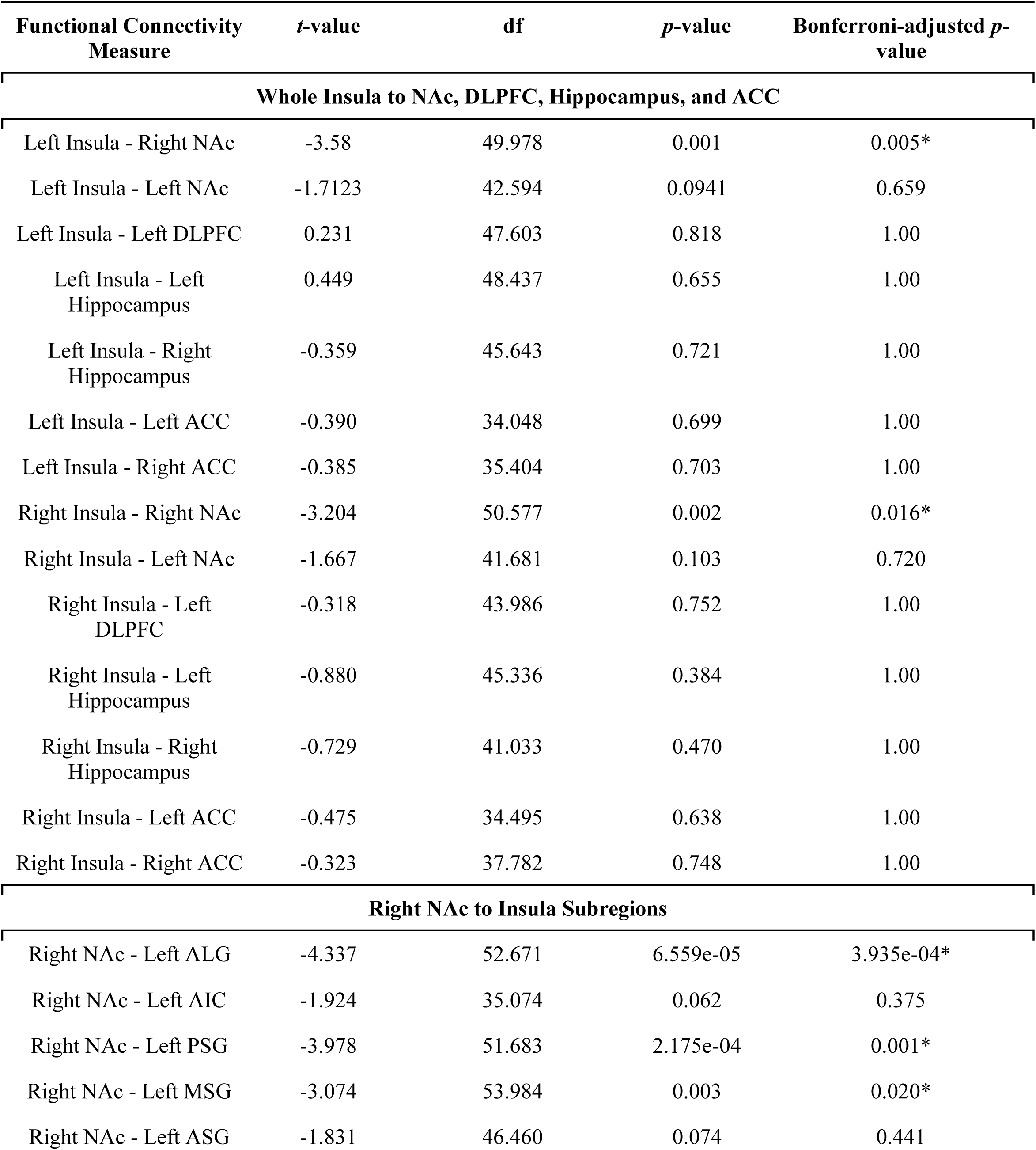
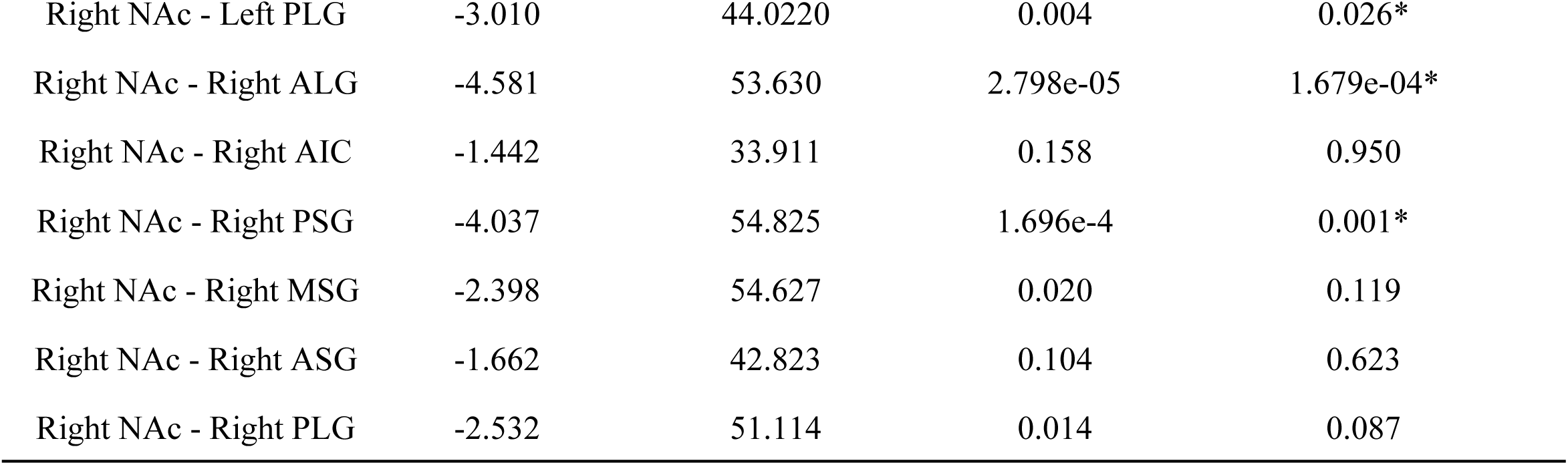
Functional connectivity was computed between the left and right whole insula and the left/right NAc, hippocampus, and ACC, in addition to the stimulation target (left DLPFC). Left and right insula functional connectivity to the right NAc was significantly lower in responders compared to nonresponders. Subsequently, functional connectivity between the right NAc and 12 insula sub-regions was computed. Functional connectivity with the right NAc was significantly lower in responders for the following insula sub-regions: left ALG, left PSG, left MSG, left PLG, right ALG, and right PSG. Abbreviations: anterior long gyrus (ALG), anterior inferior cortex (AIC), posterior short gyrus (PSG), middle short gyrus (MSG), anterior short gyrus (ASG) and posterior long gyrus (PLG).

| Functional Connectivity Measure | *t*-value | df | *p*-value | Bonferroni-adjusted *p*-value |
| --- | --- | --- | --- | --- |
| **Whole Insula to NAc, DLPFC, Hippocampus, and ACC** |  |  |  |  |
| Left Insula - Right NAc | -3.58 | 49.978 | 0.001 | 0.005* |
| Left Insula - Left NAc | -1.7123 | 42.594 | 0.0941 | 0.659 |
| Left Insula - Left DLPFC | 0.231 | 47.603 | 0.818 | 1.00 |
| Left Insula - Left Hippocampus | 0.449 | 48.437 | 0.655 | 1.00 |
| Left Insula - Right Hippocampus | -0.359 | 45.643 | 0.721 | 1.00 |
| Left Insula - Left ACC | -0.390 | 34.048 | 0.699 | 1.00 |
| Left Insula - Right ACC | -0.385 | 35.404 | 0.703 | 1.00 |
| Right Insula - Right NAc | -3.204 | 50.577 | 0.002 | 0.016* |
| Right Insula - Left NAc | -1.667 | 41.681 | 0.103 | 0.720 |
| Right Insula - Left DLPFC | -0.318 | 43.986 | 0.752 | 1.00 |
| Right Insula - Left Hippocampus | -0.880 | 45.336 | 0.384 | 1.00 |
| Right Insula - Right Hippocampus | -0.729 | 41.033 | 0.470 | 1.00 |
| Right Insula - Left ACC | -0.475 | 34.495 | 0.638 | 1.00 |
| Right Insula - Right ACC | -0.323 | 37.782 | 0.748 | 1.00 |
| **Right NAc to Insula Subregions** |  |  |  |  |
| Right NAc - Left ALG | -4.337 | 52.671 | 6.559e-05 | 3.935e-04* |
| Right NAc - Left AIC | -1.924 | 35.074 | 0.062 | 0.375 |
| Right NAc - Left PSG | -3.978 | 51.683 | 2.175e-04 | 0.001* |
| Right NAc - Left MSG | -3.074 | 53.984 | 0.003 | 0.020* |
| Right NAc - Left ASG | -1.831 | 46.460 | 0.074 | 0.441 |
| Responder / Nonresponder Smoking |  |  |  |  |
| Right NAc - Left PLG | -3.010 | 44.0220 | 0.004 | 0.026* |
| Right NAc - Right ALG | -4.581 | 53.630 | 2.798e-05 | 1.679e-04* |
| Right NAc - Right AIC | -1.442 | 33.911 | 0.158 | 0.950 |
| Right NAc - Right PSG | -4.037 | 54.825 | 1.696e-4 | 0.001* |
| Right NAc - Right MSG | -2.398 | 54.627 | 0.020 | 0.119 |
| Right NAc - Right ASG | -1.662 | 42.823 | 0.104 | 0.623 |
| Right NAc - Right PLG | -2.532 | 51.114 | 0.014 | 0.087 |

Based on this finding, functional connectivity was then computed between the right NAc and 12 insula subregions. The results suggested that functional connectivity was significantly lower in responders between the right NAc and the following insula subregions: left anterior long gyrus (Cohen’s *d* = −1.091, 95% CI = −1.6778 - 0.503); left posterior short gyrus (Cohen’s *d* = - 1.011, 95% CI = −1.5932 −0.428); left middle short gyrus (Cohen’s *d* = −0.760, 95% CI = −1.329 - 0.1920); left posterior long gyrus (Cohen’s *d* = −0.817, 95% CI = −1.385 −0.243); right anterior long gyrus (Cohen’s *d* = −1.139, 95% CI = −1.729 - 0.548); and right posterior short gyrus (Cohen’s *d* = −0.981; 95% CI = −1.561 −0.401). There were no significant differences for the remaining Right NAc – insula subregion functional connectivity measures (corrected *p*-values > .05; see **Table 4** and **Figure 9**).

**Figure 9.**
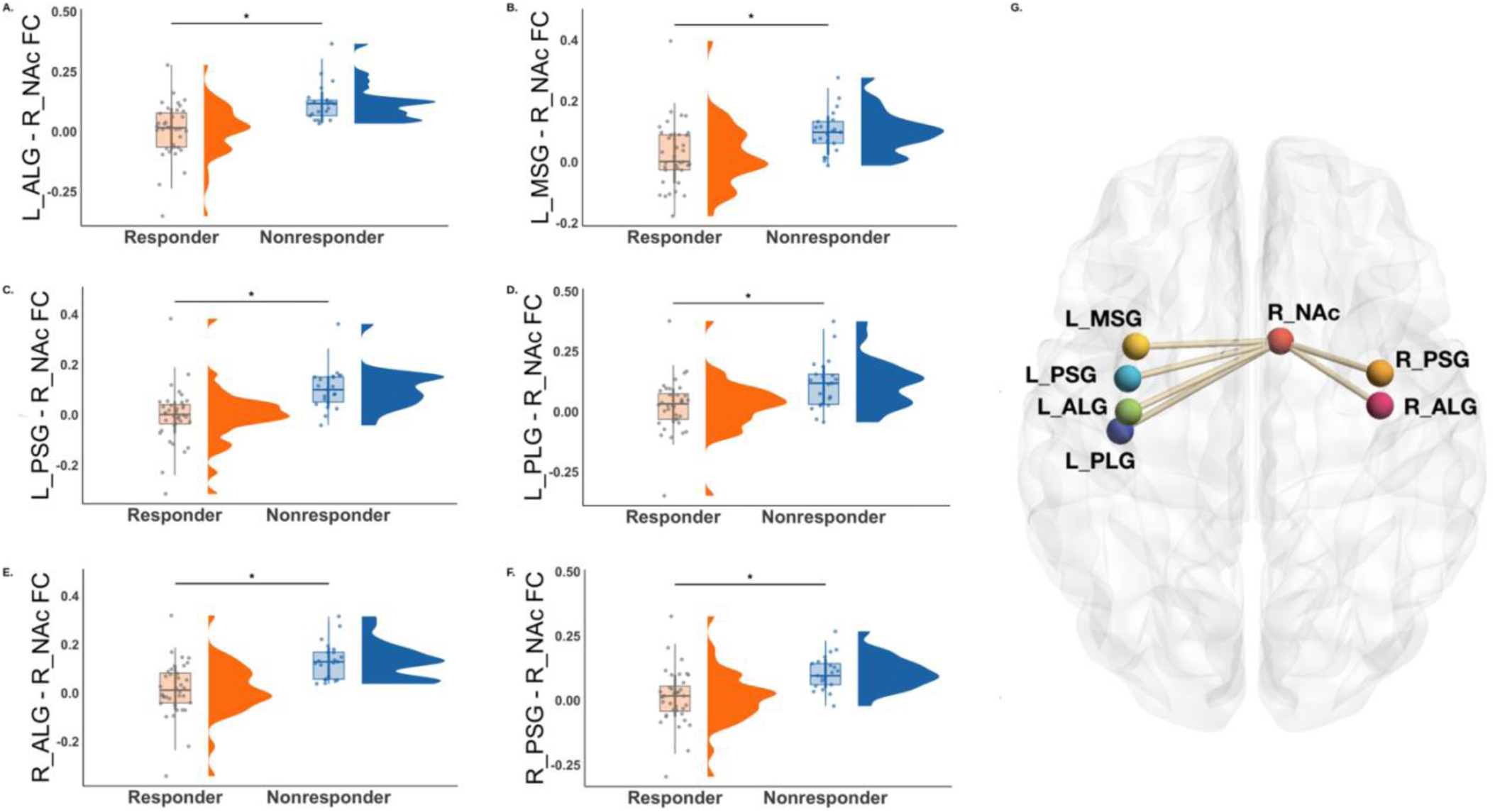
Functional connectivity (FC) was calculated between the right NAc and 12 insula sub-regions. Responders (orange), compared to nonresponders (blue), displayed reduced functional connectivity between the right NAc and 6 insula-subregions including the left anterior long gyrus (panel A), left middle short gyrus (panel B), left posterior short gyrus (panel C), left posterior long gyrus (panel D), right anterior long gyrus (panel E), and right posterior short gyrus (F). Panel G is a visualization of the NAc-insula subregion connectivity patterns that were significantly lower in responders compared to the nonresponders. \**p* < 0.05.

## Discussion

The goal of this study was to systematically test whether smoking-relevant behavioral, demographic, and neurobiological variables are associated with rTMS response in adults with TUD. rTMS to the left DLPFC, but not a control brain region (V5) led to significant reductions in self-reported cigarette craving following 12 hours of abstinence. Of the participants who received rTMS to DLPFC, 38 experienced a reduction in cigarette craving (responders) and 22 had no change in cigarette craving or experienced an increase (nonresponders). Comparisons of behavioral and demographic variables indicated that responders used significantly more cigarettes per day and experienced greater nicotine withdrawal symptoms and more cigarette cravings, suggesting that rTMS is more effective in heavier smokers who experience more severe psychological and physiological symptoms during abstinence. Given the retrospective nature of the analysis, floor effects in non-responders or a regression to the mean cannot be ruled out as alternative explanations for these effects. However, these concerns are partly mitigated by the lack of significant difference in pre-rTMS craving levels between DLPFC and V5 appointments, the use of pre-registration to identify these predictors *a priori*, and previous studies that have reported similar effects (X Li et al., 2013; Addicott et al., 2015).

The two groups did not differ significantly in age, depression symptoms, anxiety symptoms, or nicotine dependence, which in part may be because psychiatric disorders such as current major depressive disorder or generalized anxiety disorder were exclusionary in this study. As such, any depression or anxiety symptoms were sub-clinical and may have had sufficient range to influence rTMS efficacy or be detected by our statistical models. Additionally, small effect sizes were observed for rTMS coil-to-scalp distance, coil tilt, and coil rotation, indicating that small deviations in coil position throughout treatment are unrelated to rTMS response in this dataset. Average aMT was lower in responders and displayed a moderate effect size, suggesting that differences in cortical excitability may be a representation of brain functional states that are associated with response to rTMS. Similarly, past substance use and treatment for substance use disorders generated small effect sizes, which indicates that rTMS may be a feasible treatment option for individuals with varying personal substance use histories.

The pre-registered fMRI analyses were designed to test neurobiologically plausible mechanisms that may influence reductions in cigarette craving following rTMS to the DLPFC based on previous literature. Contrary to past work indicating that DLPFC functional connectivity is associated with rTMS response in substance use disorders (X. Li et al., 2017; Su et al., 2020), there were no significant differences in DLPFC-OMPFC nor DLPFC-IPL functional connectivity between responders and nonresponders. An exploratory whole-brain analysis reinforced this finding, suggesting that there were no significant differences in left DLPFC whole-brain functional connectivity between responders and nonresponders.

In previous literature, associations have been reported between insula structure/function and tobacco use topography, TUD severity, and smoking cessation outcomes (Abdolahi et al., 2015; Addicott et al., 2015; Carroll et al., 2015; Fedota et al., 2018; Gaznick et al., 2014; Ghahremani et al., 2021; Gilman et al., 2018; Janes et al., 2010, 2015; T. Jordan et al., 2024; Joutsa et al., 2022; Y. Li et al., 2017; Morales et al., 2014; Naqvi et al., 2007; Stoeckel et al., 2016; Suner-Soler et al., 2012; Wilcox et al., 2017; Zanchi et al., 2015; Zhou et al., 2017). In contrast to our hypotheses, insula seed-based connectivity for the right/left whole insula and 12 insula sub-regions did not differ significantly between responders and nonresponders. One possible explanation is that differences between responders and nonresponders were not sufficiently robust to survive voxel wise whole-brain correction. Other possibilities are that functional connectivity differences may emerge more consistently following multiple rTMS sessions rather than single session rTMS used in the current study, or a more stringent definition of “response” is required to detect differences between responders and nonresponders.

However, an alternative explanation is that our planned definition of “responders” was not optimal. In contrast to the pre-registered insula seed-based connectivity results, when exploring insula connectivity differences with “responders” defined as the ∼1/3 of participants who experienced the greatest reduction in craving after rTMS (*n* = 20), the results indicated that nonresponders had significantly greater functional connectivity between the left insula, occipital pole, and precuneus. Similarly, nonresponders had significantly greater right insula functional connectivity to the occipital pole. Although insula-based whole-brain connectivity differences were not detected using the original “responder” definition, these results indicate that functional connectivity differences are more prevalent in participants who experienced the greatest benefit from rTMS. A plausible interpretation is that nonresponders experience an abstinence-state characterized by increased coupling between brain regions responsible for interoceptive salience signals (insula), self-referential processing (precuneus), and reactivity to emotion-inducing visual stimuli such as drug cues (occipital pole) that is less modifiable by top-down DLPFC stimulation (Amedi et al., 2004; Dadario and Sughrue, 2023; McCalley et al., 2025). Subsequent exploratory ROI-ROI analyses indicated that responders had reduced functional connectivity between the right NAc and the left/right whole insula and insula subregions including the left posterior short gyrus, left middle short gyrus, left posterior long gyrus, right anterior long gyrus, and right posterior short gyrus. This may represent reduced integration between salience/interoceptive systems and reward circuitry (Uddin et al., 2017; Balfour, 2004), such that craving-related functional connectivity patterns are more modifiable with DLPFC stimulation. Importantly, this effect was observed for multiple insula subregions spanning the anterior, middle, and posterior insula. This suggests that distinct insula subregions involved in cognitive control (dorsal anterior insula), affect (ventral anterior insula), and sensorimotor processes (posterior insula) are interacting with reward circuitry (NAc) in contributing to rTMS response (Faillenot et al., 2017; Menon & Uddin, 2010; Uddin et al., 2014; Uddin, 2015; Uddin et al., 2017).

It is important to note adjustments from the pre-registered hypotheses and analysis plan. The original plan was to analyze data from *N* = 45 participants, but the final sample included *N* = 60, and therefore a larger dataset was analyzed to increase power. Additionally, the pre-registered plan did not specify differences in effect size calculations between continuous and categorical variables and did not include computing Fisher’s exact test for categorical variables. Another adjustment was that the pre-registered analysis plan stated that the IPL ROI would be an 8mm sphere, but it was changed to a 5mm sphere to match the OMPFC ROI. Finally, although significant effects were detected in the exploratory analyses and the effects survived multiple comparison correction, they were not part of the pre-registered analysis plan. As such, they should be interpreted cautiously and be viewed as hypothesis-generating rather than hypothesis-confirming.

### Limitations and Future Directions

Despite a rigorous, pre-registered design that explored variables that were predicted to be associated with rTMS response, the current study had limitations that can be used to guide future research. The main limitation of the current study is the use of single-session rTMS. In order to generate clinically meaningful and lasting effects, multiple rTMS sessions are required (Apostol et al., 2026). That said, single-session rTMS may be a useful tool for parameter optimization or to test hypotheses about rTMS response. For instance, (Zangen et al., 2021) reported that responding to the first session of a multi-session rTMS treatment course for smoking cessation was associated with responding to the overall treatment course. Therefore, it is possible that the factors that are associated with responding to single-session rTMS would predict responding to a multi-session rTMS course, but future work is required to answer this question. Additionally, it has not been empirically demonstrated that the rTMS targeting approaches used here confer advantages over other targeting approaches (e.g., Beam F3 method; Beam et al., 2009). While neuronavigation and functional connectivity-based targets are associated with coil placement reproducibility (Caulfield et al., 2022), it is unclear if it generates improved reductions in cigarette craving or other clinically relevant variables. Future studies should focus on determining the relative efficacy of fMRI-based rTMS targeting compared to conventional rTMS targeting approaches in the context of smoking cessation treatment. With rTMS coil positioning and localization, there may be tradeoffs between cost, time, and efficacy, and exploring these factors in the context of addiction treatment should be a focus of future work. Future studies should focus on the efficacy of other rTMS parameters in the context of smoking cessation. For instance, it is unclear if there are different predictors of rTMS response when using 10 Hz rTMS versus intermittent theta burst stimulation (iTBS). Finally, the results reported in this study could be used to inform treatment stratification in future randomized clinical trials testing the efficacy of accelerated rTMS for smoking cessation (Cole et al., 2020, 2022).

The sample in the current study is limited by the exclusion of other substance use disorders or psychiatric disorders. There is substantial overlap between TUD, other substance use (Hughes, 1999), and psychiatric disorders, and exploring the effects of rTMS in individuals with comorbidities is a critical future direction. Additionally, the study design is limited by the reliance of self-report nicotine withdrawal and cigarette craving measures as the primary outcome measures of treatment efficacy. Although these measures were biochemically verified using CO and cotinine as biomarkers, additional steps could have been taken to increase rigor. For instance, the use of fitness tracker wristbands to measure physiological states could corroborate self-report findings. Finally, while there was a minimum 24-hour washout period between rTMS appointments, there may have been crossover effects (although, our previous results indicate this is not the case; Apostol et al., 2023; Petersen et al., 2025).

The pre-registered analysis plan included *a priori* specified analyses. While this is an advantage in increasing experimental and statistical rigor, this approach is limited in the constrained ROIs and analyses that were performed. For instance, defining “response” as a continuous variable rather than dichotomizing participants into “responder” and “nonresponder” categories is a viable alternative approach. Alternatively, data-driven approaches examining whole-brain functional connectivity patterns or other fMRI analysis modalities should be explored in future work, such as dynamic functional connectivity (Nomi et al., 2016) or brain signal variability (T. Jordan et al., 2024; Nomi et al., 2017), as they may better detect differences between rTMS responders and nonresponders. Finally, based on data availability and statistical considerations, adjustments were made to the pre-registered analysis plan for this study.

## Conclusion

In summary, the goal of this study was to perform a within-subjects, single-blind, active-controlled, randomized, crossover experiment designed to identify whether rTMS responders and nonresponders display significant differences in behavioral, demographic, and neuroimaging variables. rTMS to the DLPFC led to significant reductions in cigarette craving. Although responders reported greater levels of cigarette consumption, nicotine withdrawal, and cigarette craving, they exhibited reduced functional connectivity between the insula and regions involved in reward (NAc), self-referential processing (precuneus), and visual cue processing (occipital pole). One interpretation is that responders experienced a more intense nicotine abstinence state that is embodied by reduced functional connectivity between regions implicated in salience and reward processing. Reduced functional connectivity in these circuits, in turn, may be more modifiable with rTMS applied to the DLPFC. This study and future investigations that aim to identify predictors of rTMS will lead to a better understanding of rTMS as an addiction treatment, more precise treatment stratification, personalized stimulation protocols, and better smoking cessation outcomes.

## Data Availability

Data are available from the authors upon reasonable request.

